# Machine learning-based short-term forecasting of COVID-19 hospital admissions using routine hospital patient data

**DOI:** 10.1101/2025.05.21.25328056

**Authors:** Martin S. Wohlfender, Judith A. Bouman, Olga Endrich, Alban Ramette, Alexander B. Leichtle, Guido Beldi, Christian L. Althaus, Julien Riou

## Abstract

During the COVID-19 pandemic, the field of infectious disease modeling advanced rapidly, with forecasting tools developed to track trends in transmission dynamics and anticipate potential shortages of critical resources such as hospital capacity. In this study, we compared short-term forecasting approaches for COVID-19 hospital admissions that generate forecasts one to five weeks ahead, using retrospective electronic health records. We extracted different features (e.g., daily emergency department visits) from an individual-level patient dataset covering six hospitals located in the region of Bern, Switzerland from February 2020 to June 2023. We then applied five methods – last-observation carried forward (baseline), linear regression, XGBoost and two types of neural networks – to time series using a leave-future-out training scheme with multiple cutting points and optimized hyperparameters. Performance was evaluated using the root mean square error between forecasts and observations. Generally, we found that XGBoost outperformed the other methods in predicting future hospital admissions. Our results also show that adding features such as the number of hospital admissions with fever and augmenting hospital data with measurements of viral concentration in wastewater improves forecast accuracy. This study offers a thorough and systematic comparison of methods applicable to routine hospital data for real-time epidemic forecasting. With the increasing availability and volume of electronic health records, improved forecasting methods will contribute to more precise and timely information during epidemic waves of COVID-19 and other respiratory viruses, thereby strengthening evidence-based public health decision-making.

## 1. Introduction

The COVID-19 pandemic has highlighted the need for reliable infectious disease monitoring and forecasting systems. As SARS-CoV-2 spread globally, researchers, healthcare professionals, public health authorities, and governments undertook extensive efforts to mitigate its impact and control transmission dynamics. A key priority was to ensure that hospital capacity, particularly in intensive care units, was not exceeded. Whenever hospital capacities were exceeded, hospitals were forced to implement crisis care standards, including treatment protocol classifications that prioritized patients with the highest probability of survival, often leading to delayed or reduced care for other patients. This also resulted in the postponement of elective procedures, in increased stress and burnout among healthcare workers, and in higher mortality rates due to limited access to critical resources such as ICU beds and ventilators (Anderegg et al., 2022; Didriksson et al., 2022). In such situations, short-term forecasts aimed at anticipating new hospital admissions a few weeks in advance can be invaluable for public health decision makers and hospital management.

Researchers worldwide have applied numerous approaches to forecast the spread and impact of SARS-CoV-2 in different settings based on various types of data. Due to increasing digitization, substantially more data reflecting various aspects of the pathogen were collected during the COVID-19 pandemic compared to the historic major infectious disease outbreaks. Examples of such data are wearable or smartphone sensor data (Grantz et al., 2020), viral genome sequences (Shu and McCauley, 2017; Furuse, 2021; CDC, 2024; Hodcroft et al., 2025), viral load measurements in wastewater (Morvan et al., 2022; Jahn et al., 2022), and electronic health records from hospitals and medical practices (Qian et al., 2021). A wide variety of methods have been used to produce forecasts, including mechanistic models (e.g., deterministic or stochastic compartmental models, agent-based models), statistical time series models (e.g., ARIMA, exponential smoothing, regression) and machine learning methods (e.g., tree-based models, neural networks) (Kraemer et al., 2025). In both the United States and Europe, groups of scientists developed standardized forecasting pipelines for COVID-19 cases, hospital admissions, and deaths in different geographic regions (Cramer et al., 2021; ECDC, 2021, 2023). This allowed the combination of multiple models from different groups into an ensemble forecast with a single cone of uncertainty. Hospital capacity is an important indicator when planning public health interventions during major outbreak of an infectious disease. Therefore, models that provide estimates of expected admissions to hospitals on a national or local level in the coming weeks can be of great benefit for taking decisions on the introduction of public health measures. One approach has been to systematically test for infection with SARS-CoV-2 all patients hospitalized for elective procedures, which outperformed state-based data in predicting the local clinical burden(Covello et al., 2021). Furthermore, more detailed hospital data like ICU admission and discharge or ambulance service and emergency unit notes have been used for predicting COVID-19-related hospital admissions within a region (Qian et al., 2021; Ferté et al., 2022). Augmenting data extracted from electronic health records with exogenous variables like weather or mobility data also lead to more accurate forecasts of local COVID-19 related hospital admissions compared to using hospital data alone Ferté et al. (2022); Zhang et al. (2022); Klein et al. (2023).

In this study, we compared the performance of different machine learning models to forecast the number of COVID-19 hospital admissions based on routinely collected electronic health records (EHR) and wastewater data. We hypothesized that quantities such as the occupancy of a hospital’s emergency ward, vital signs of hospital patients such as fever, or measurements of viral load in wastewater have high predictive power for short-term forecasting of COVID-19-related hospital admissions and lead to more accurate predictions than relying on the number of hospital admissions in the previous days alone. First, we extracted candidate variables that could have high predictive power for the spread of SARS-CoV-2 from a large individual patient-level EHR dataset from six hospitals in the Bern region, Switzerland, in the period from February 2020 to June 2023. Second, we trained different machine learning models with different combinations of features on the data to forecast the number of COVID-19 hospital admissions up to five weeks in advance. Third, we evaluated the performance of the models in comparison to a baseline model across different forecasting setups.

## 2. Data and Methods

### 2.1. Forecasting setup

This study aimed to validate and compare methods for forecasting the weekly number of COVID-19 hospital admissions up to five weeks in advance, using routinely collected hospital data from the previous days. As a case study, we drew on electronic health records (EHR) data from six hospitals belonging to the Insel Gruppe network, all located in the region of Bern, Switzerland, collected from 25 February 2020, the day the first COVID-19 case was detected in Switzerland (FOPH, Federal Office of Public Health, 2020), and 30 June 2023 (full study period). We adopted a retrospective approach by applying five forecasting models to historical time series data – where outcomes are already known – enabling a comparison of the performance of each method. We employed a leave-future-out strategy incorporating 12 separate test datasets, each covering a period of two to four months. We selected the 12 cut-off points based on peaks and valleys of the daily time series of COVID-19 hospital admissions in the next seven days (Supplementary Figure S1 A and B of Appendix A). The training datasets contained all data collected before the respective cut-off point (Supplementary Figure S1 C and D of Appendix A). The target week was defined as the sliding seven-day window for which hospital admissions were forecast with the trained models (Figure 1). We systematically varied the forecasting horizon *k* (i.e., the gap between the last day of observed data and the start of the target week) and the lookback window *p* (i.e., the number of past days of data included in the model). More formally, placing ourselves at time *t*, we used data from days *{t−p, t−p*+1*, … , t−*1*}* to forecast the number of COVID-19 hospital admissions during the target week *{t* + *k, t* + *k* + 1*, … , t* + *k* + 6*}*. In our analysis, we used the following sets *k* = *{*0, 7, 14, 21, 28*}* and *p* = *{*7, 14, 21, 28, 35*}*. We did not consider forecasting horizons beyond five weeks as transmission dynamics – and the many factors that influence them – are likely to shift rapidly within that period (Holmdahl and Buckee, 2020).

**Figure 1:**
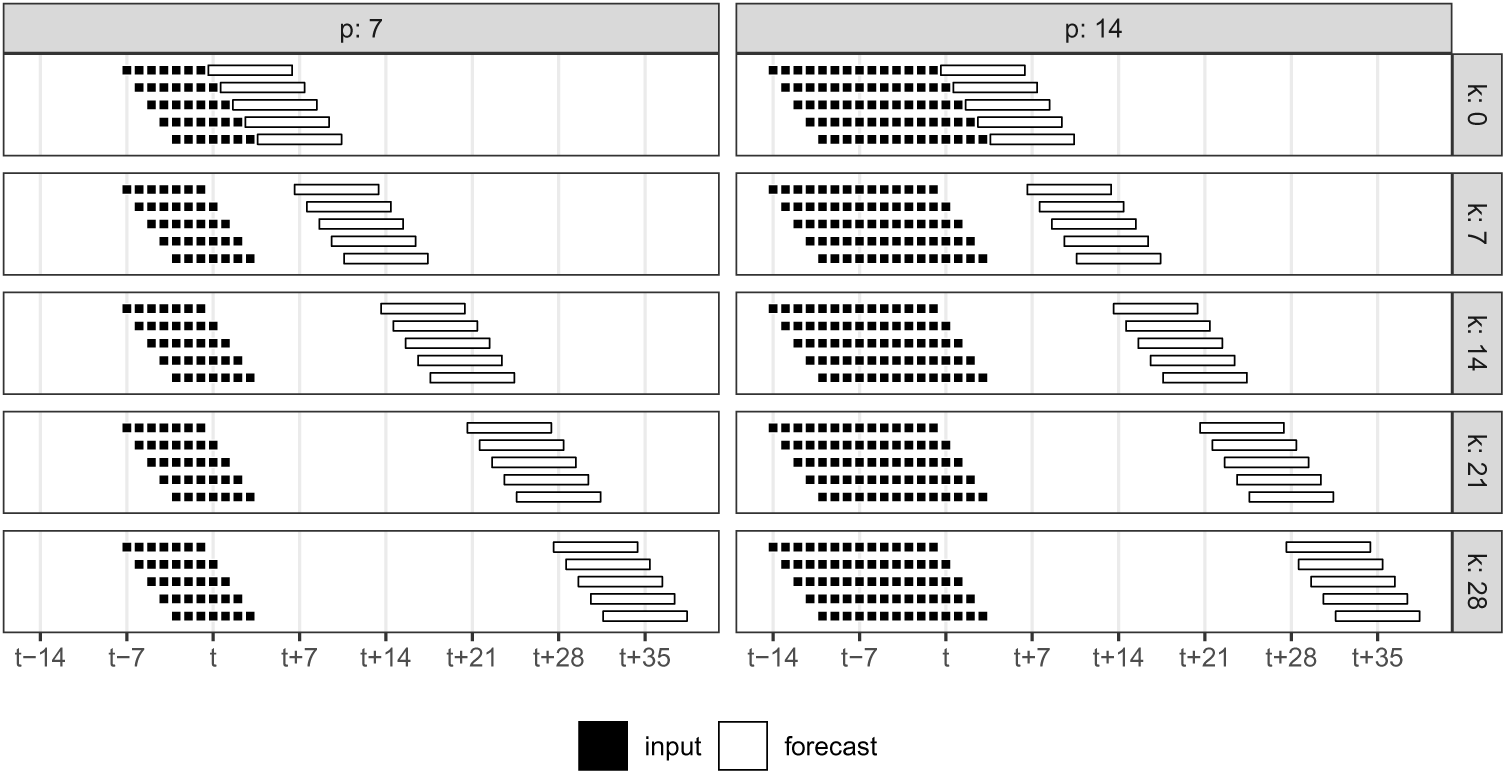
Forecasting setups. The forecasting horizon *k* corresponds to the gap between the last day of observed data and the start of the target week. The lookback window *p* is the number of past days of data included in the model. Day *t* corresponds to the start of a testing period.

### 2.2. Electronic health records data

We obtained individual-level electronic health records (EHR) from the Insel Gruppe hospital network (inselgruppe.ch) in the canton of Bern, Switzerland. During the study period, this hospital network comprised Bern University Hospital, which is one of the five first-level university general hospitals of Switzerland, as well as five other hospitals (Aarberg, Belp, Münsingen, Riggisberg and Tiefenau) that are second-level general hospitals (FOPH, Federal Office of Public Health, 2023). In 2023, about 57,000 inpatients and 900,000 outpatients were treated at Insel Gruppe hospitals (Inselgruppe, 2023). The full dataset covers the period from 1 January 2014 to 30 June 2023. It contains personal information about patients (e.g., age and sex), details of their hospital stay (e.g., dates of admission and discharge, hospital ward), as well as various clinical and laboratory measurements (e.g., body temperature, blood pressure, C-reactive protein [CRP] concentration). In addition, diagnoses of inpatients were recorded using ICD10 codes (WHO, 2019). These codes were assigned after discharge by trained medical coders based on the clinical documentation – including medical doctors’ notes, laboratory results, and imaging reports. This process is primarily done for administrative and financial purposes, but can be leveraged for epidemiological monitoring (Demont et al.).

### 2.3. Wastewater data

In addition to EHR, we included measurements of the concentration of SARS-CoV-2 RNA in wastewater. We used wastewater samples collected daily at the Sensetal Laupen treatment plant between 16 November 2021 and 30 June 2023 (partial study period) as part of a wastewater surveillance program coordinated by the Swiss Federal Institute of Aquatic Science and Technology (Eawag) and the Swiss Federal Office of Public Health (FOPH) (Eawag, 2021). As process control identified a possible underestimation by approximately 30% of the SARS-CoV-2 viral load in wastewater during summer 2022, there is a five-week interruption in the data between 13 July and 16 August 2022. This plant covers approximately 62,000 people living in an area west of the city of Bern, overlapping with the of the catchment area of the Insel Gruppe hospital network. Samples were stored on-site at 4*°*C and transported in batches to a laboratory for concentration, nucleic acid extraction, and quantification using qPCR. Further details on the wastewater sample laboratory procedures are available elsewhere (Huisman et al.).

### 2.4. Data processing

These raw data were processed to create 25 daily time series (Figure 2) in three steps. First, we identified COVID-19-related hospital admissions using the ICD10 code U07.1 (“*COVID-19, virus identified* ” WHO (2019)) and created daily time series. We then smoothed this time series using a seven-day moving sum to reduce day to day fluctuations and focus on the actual trend of the time series. The entry at day *t* of the smoothed daily time series corresponds to the total number of COVID-19-related hospital admissions during days *t* to *t* + 6. This time series was used as the target variable in all models. Furthermore, we stratified COVID-19 hospital admissions into five age groups: Ages *≤* 4, 5 *−* 14, 15 *−* 29, 30 *−* 64 and *≥* 65 years. Second, we created several other daily time series to be used as features in the models. These included the number of patients seeking care at the emergency department of Bern University Hospital, from both patients that were admitted to another hospital ward afterwards as well as patients discharged directly. Next, we identified the daily number of hospital admissions including a diagnosis belonging to one of five ICD10 chapters (R, I, E, J or Z), belonging to one of five ICD10 categories (E87, J12, J96, I10, N18) or including one of five specific ICD10 codes (J12.8, I10.90, J96.00, Z22.8, or B33.8) (details about each code are available in Table 1). These chapters, categories and codes were selected on the basis of the frequency with which they appear together with the ICD10 code U07.1 in patients’ diagnoses. We also determined the daily number of inpatients admitted to hospital with fever (*≥* 38.5 ^◦^C) and the daily number of inpatients admitted to hospital with a high CRP concentration (*≥* 50 mg*/*l). Third, we processed SARS-CoV-2 wastewater concentration data by 1) normalizing measurement using the flow of wastewater on the sampling day as in common practice (Huisman et al.), and 2) filling missing values using linear interpolation. Note that wastewater data were only available for a shorter time period, referred to as the partial study period in the following. From these 25 times series, we built 9 feature sets for the full study period referred to by letters A to I (without wastewater) and 3 additional feature sets for only the partial study period referred to by letters J to L (with wastewater) (Tables 2 and 3).

**Figure 2:**
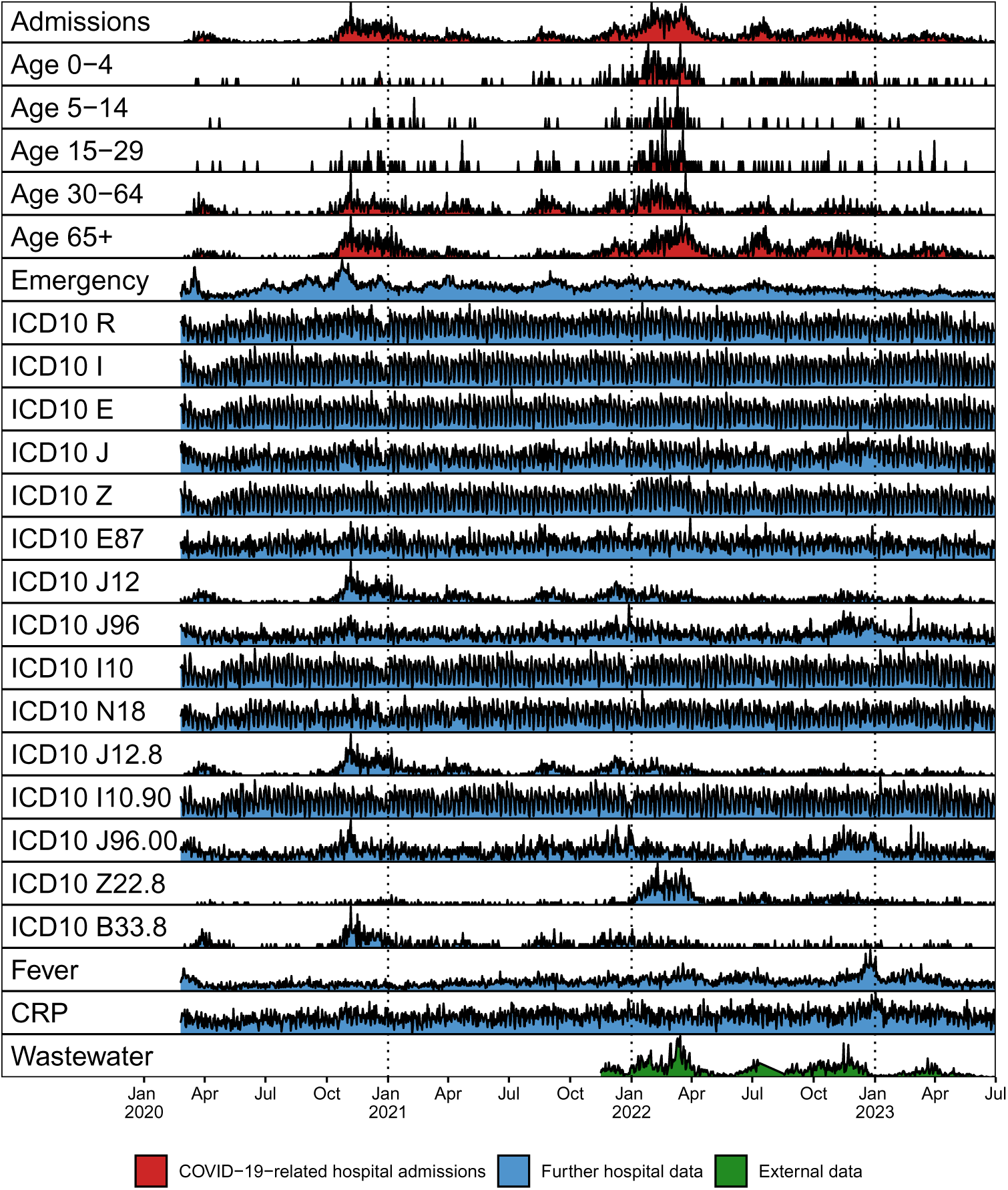
Temporal profile of model variables. Variables are extracted from electronic health records and wastewater data. All time series are normalized and on a daily level.

**Table 1:** Summary characteristics of model variables. Abbreviation, definition and use in feature sets of all input variables extracted from electronic health records (EHR) and wastewater data. Sum, mean, minimum and maximum are taken across all days of the full study period for the EHR data and across all days of the partial study period for the wastewater data. The unit of the variables extracted from EHR data is the number of new hospital admissions fulfilling a certain criterion per day. The unit of the viral load in wastewater samples is the number of SARS-CoV-2 RNA copies per 100, 000 people in the collection area and day.

| Variable | Details | Feature sets | Sum | Daily Mean | Daily Min | Daily Max |
| --- | --- | --- | --- | --- | --- | --- |
| Admissions | COVID-19-related hospital admissions (any age) | A-I and K-L | 6,038 | 4.9 | 0 | 31 |
| Age 0-4 | COVID-19-related hospital admissions (age 0-4) | B | 389 | 0.3 | 0 | 6 |
| Age 5-14 | COVID-19-related hospital admissions (age 5-14) | B | 108 | 0.1 | 0 | 4 |
| Age 15-29 | COVID-19-related hospital admissions (age 15-29) | B | 220 | 0.2 | 0 | 4 |
| Age 30-64 | COVID-19-related hospital admissions (age 30-64) | B | 1,840 | 1.5 | 0 | 13 |
| Age 65+ | COVID-19-related hospital admissions (age 65+) | B | 3,481 | 2.8 | 0 | 18 |
| Emergency | Patients seeking treatment at Bern University Hospital Emergency Department | C, I and L | 200,895 | 164.4 | 74 | 348 |
| ICD10 R | Symptoms, signs and abnormal clinical and laboratory findings, not elsewhere classified | D | 68,246 | 55.8 | 18 | 99 |
| ICD10 I | Diseases of the circulatory system | D | 97,842 | 80.1 | 22 | 144 |
| ICD10 E | Endocrine, nutritional and metabolic diseases | D | 86,092 | 70.5 | 17 | 132 |
| ICD10 J | Diseases of the respiratory system | D | 39,121 | 32.0 | 7 | 65 |
| ICD10 Z | Factors influencing health status and contact with health services | D | 102,570 | 83.9 | 20 | 165 |
| ICD10 E87 | Other disorders of fluid, electrolyte and acid-base balance | E | 19,577 | 16.0 | 1 | 39 |
| ICD10 J12 | Viral pneumonia, not elsewhere classified | E | 2,745 | 2.2 | 0 | 21 |
| ICD10 J96 | Respiratory failure, not elsewhere classified | E | 9,877 | 8.1 | 0 | 27 |
| ICD10 I10 | Essential (primary) hypertension | E | 44,497 | 36.4 | 7 | 72 |
| ICD10 N18 | Chronic kidney disease | E | 27,847 | 22.8 | 2 | 49 |
| ICD10 J12.8 | Other viral pneumonia | F, I and L | 2,426 | 2.0 | 0 | 21 |
| ICD10 I10.90 | Essential hypertension, unspecified without indication of hypertensive crisis | F, I and L | 40,909 | 33.5 | 6 | 69 |
| ICD10 J96.00 | Acute respiratory failure, not elsewhere classified | F, I and L | 5,361 | 4.4 | 0 | 17 |
| ICD10 Z22.8 | Carrier of other infectious diseases | F, I and L | 1,344 | 1.1 | 0 | 17 |
| ICD10 B33.8 | Other specified viral diseases | F, I and L | 808 | 0.7 | 0 | 11 |
| Fever | Highest body temperature measurement on day of admission at least 38.5 degrees Celsius | G, I and L | 16,589 | 13.6 | 1 | 56 |
| CRP | Highest CRP concentration measurement on day of admission at least 50 mg/l | H, I and L | 28,726 | 23.5 | 3 | 50 |
| Wastewater | SARS-CoV-2 RNA copies per 100,000 people living in the collection area of the Sensetal Laupen wastewater treatment plant and day | J, K and L | $4.0 \times 10^{15}$ | $6.8 \times 10^{12}$ | $2.3 \times 10^{10}$ | $3.7 \times 10^{13}$ |

**Table 2:** Summary of model performance for the full study period (25 February 2020 to 30 June 2023). For each model and feature set, the summary score was computed as the geometric mean of the ratios of the root mean square error (RMSE) over the baseline RMSE across all combinations of forecasting horizon *k*, lookback window *p*, and train-test split. The proportion of forecasts where the RMSE is lower than the baseline is shown in parentheses.

| Feature set | LOCF | LR | RNN | LSTM | XGBoost |
| --- | --- | --- | --- | --- | --- |
| A (COVID-19-related hospital admissions) | 1 | 1.08 (48 %) | 0.91 (69 %) | 0.90 (68 %) | 0.88 (76 %) |
| B (A + COVID-19-related hospital admissions by age group) | 1 | 1.69 (19 %) | 0.96 (58 %) | 1.01 (55 %) | 0.87 (75 %) |
| C (A + emergency) | 1 | 1.17 (51 %) | 0.89 (70 %) | 0.89 (58 %) | 0.90 (69 %) |
| D (A + ICD10 chapters) | 1 | 1.51 (28 %) | 0.98 (57 %) | 0.93 (56 %) | 0.86 (77 %) |
| E (A + ICD10 categories) | 1 | 1.68 (16 %) | 1.00 (55 %) | 0.95 (58 %) | 0.87 (75 %) |
| F (A + ICD10 codes) | 1 | 1.82 (17 %) | 1.02 (46 %) | 1.02 (48 %) | 0.89 (75 %) |
| G (A + fever) | 1 | 1.16 (39 %) | 0.98 (56 %) | 0.91 (61 %) | 0.87 (75 %) |
| H (A + CRP) | 1 | 1.28 (37 %) | 0.98 (53 %) | 0.96 (54 %) | 0.87 (76 %) |
| I (A + emergency + ICD10 codes + fever + CRP) | 1 | 2.02 (12 %) | 1.12 (37 %) | 1.01 (46 %) | 0.91 (71 %) |

**Table 3:** Summary of model performance for the partial study period (16 November 2021 to 30 June 2023). For each model and feature set, the summary score was computed as the geometric mean of the ratios of the root mean square error (RMSE) over the baseline RMSE across all combinations of forecasting horizon *k*, look-back window *p*, and train-test split. The proportion of forecasts where the RMSE is lower than the baseline is shown in parentheses.

| Feature set | LOCF | LR | RNN | LSTM | XGBoost |
| --- | --- | --- | --- | --- | --- |
| A (COVID-19-related hospital admissions) | 1 | 1.45 (14 %) | 1.16 (43 %) | 1.35 (32 %) | 0.81 (78 %) |
| B (A + COVID-19-related hospital admissions by age group) | 1 | 2.26 ( 2 %) | 1.05 (53 %) | 1.39 (32 %) | 0.80 (79 %) |
| C (A + emergency) | 1 | 1.02 (45 %) | 0.83 (75 %) | 1.01 (58 %) | 0.89 (69 %) |
| D (A + ICD10 chapters) | 1 | 2.24 ( 0 %) | 1.46 (24 %) | 1.33 (34 %) | 0.78 (81 %) |
| E (A + ICD10 categories) | 1 | 3.06 ( 3 %) | 1.52 (17 %) | 1.50 (23 %) | 0.82 (67 %) |
| F (A + ICD10 codes) | 1 | 2.42 ( 0 %) | 1.32 (29 %) | 1.41 (26 %) | 0.81 (75 %) |
| G (A + fever) | 1 | 1.77 ( 4 %) | 1.19 (49 %) | 1.20 (36 %) | 0.75 (90 %) |
| H (A + CRP) | 1 | 1.73 ( 8 %) | 1.54 (15 %) | 1.51 (23 %) | 0.81 (77 %) |
| I (A + emergency + ICD10 codes + fever + CRP) | 1 | 3.50 ( 0 %) | 1.08 (60 %) | 1.23 (44 %) | 0.82 (74 %) |
| J (Wastewater) | 1 | 2.04 ( 0 %) | 1.33 (34 %) | 1.50 (30 %) | 0.74 (89 %) |
| K (A + wastewater) | 1 | 1.51 (16 %) | 1.13 (45 %) | 1.38 (32 %) | 0.73 (97 %) |
| L (A + emergency + ICD10 codes + fever + CRP + wastewater) | 1 | 3.77 ( 2 %) | 1.02 (61 %) | 1.22 (44 %) | 0.75 (86 %) |

### 2.5. Models

We applied several supervised machine learning models, each based on a different algorithm, to forecast COVID-19 hospital admissions. We selected last observation carried forward (LOCF) to serve as the baseline for performance comparison. Four models were evaluated : (1) a simple linear regression (LR) model (using base R lm function), (2) a recurrent neural network (RNN) (using Python library Keras (Chollet et al., 2015)), 3) a long short-term memory (LSTM) neural network model (Hochreiter and Schmidhuber, 1997) (using Python library Keras (Chollet et al., 2015)), and 4) a gradient boosting model (XGBoost) (XGBoost community, 2025) (using R package xgboost (Chen et al., 2024)). For both RNN and LSTM, we used a grid-search strategy to optimize the architecture of the network, the activation function and several hyperparameters (144 combinations each). For XGBoost, we evaluated 864 combinations of hyperparameters, including maximal tree depth. In all cases, the optimization of hyperparameters was based on the root mean square error (RMSE) between forecasts and observations in the test set. A complete list of all hyperparameters for all models is included in Supplementary Table S1 of Appendix A. Model forecasts of COVID-19 hospital admissions were directly taken as forecasts in the case of XGBoost, while for the RNN and LSTM models forecasts were averaged over 50 independent runs (i.e., the forecasts correspond to an ensemble mean taken sample-wise across 50 independent runs of the same model with different random seeds).

### 2.6. Evaluation of model performance

We used a summary score based on RMSE to evaluate the predictive performance of the different models to forecast COVID-19 hospital admissions in comparison to the baseline model LOCF across a range of experimental conditions. For a collection of forecasts, we first determined for each the RMSE between forecast and observed values in the respective test set. Second, we divided the obtained number by the RMSE resulting from the forecast of the baseline model LOCF in the same conditions. Finally, we aggregated these ratios into a single number by computing their geometric mean. We included a more formal definition of the summary score in Chapter 1.5 of Appendix A. This metric was computed for every combination of model and feature set, separately for the full (without wastewater) and the partial study period (with wastewater). As an additional metric for these collections of forecasts, we computed the percentage of forecasts that achieved a lower RMSE than the forecast of the baseline model LOCF with the same forecasting horizon *k*. We also computed the summary score within additional levels of stratification (e.g., for each combination of *k* and *p*) to identify which models performed best across different conditions. The summary score provided a clear and interpretable measure of performance: values below 1 indicate that on average the model forecasts considered are more accurate than the forecasts of the baseline model LOCF, while values above 1 suggest inferior performance.

### 2.7. Data and code availability

All code written in R and Python as well as some data and results files are publicly available in the GitHub repository (github.com/mwohlfender/ hospital_admission_forecasting). Due to data protection regulations we can not make the full hospital dataset publicly available, but only in aggregated form.

## 3. Results

Between 25 February 2020 and 30 June 2023, we identified 6, 038 COVID-19-related inpatient admissions, i.e. hospital stays of at least one night with ICD10 code U07.1, in 6 hospitals in the canton of Bern, Switzerland (Table 1 and Figure 2). 389 patients (6.4%) were 0 *−* 4, 108 patients (1.8%) 5 *−*14, 220 patients (3.6%) 15 *−* 29, 1840 patients (30.5%) 30 *−* 64 and 3481 patients (57.7%) were at least 65 years old. 527 of 717 COVID-19-related inpatient admissions of patients below the age of 30 occurred between 1 January 2022 and 30 June 2023. The peaks in the number of visits at the emergency ward of Bern University Hospital in mid-march 2020 and in late October 2020 reflect rapid increases in COVID-19 cases in Switzerland. The Omicron variant did not lead to a distinct increase of the number of patients seeking care at the emergency ward of Bern in December 2022 or January 2023. In autumn 2020, the trend in COVID-19-related hospital admissions coincided with that of hospital admissions with ICD10 codes J12.8 (“*Other viral pneumonia*”) and B33.8 (“*Other specified viral diseases*”). A similar pattern was observed in the first half of 2022 with ICD10 code Z22.8 (“*Carrier of other infectious diseases*”). The wastewater data showed generally similar trends as the COVID-19-related hospital admissions time series.

We combined the outputs of models with varying forecasting horizons to generate forecasts of the weekly number of COVID-19-related hospital admissions up to five weeks ahead. Examples of such forecasts are presented in Figure 3. We found that no single combination of model and feature set consistently produced the most accurate forecasts. The precision of forecasts, as measured by the RMSE between forecasts and observations in the test set, varied substantially across time periods, models, feature sets, lookback windows and forecasting horizons. Overall, forecast precision improved as more data became available for model training. The largest discrepancies between forecasts and observed values were observed during periods with rapid increases of COVID-19-related hospital admissions.

**Figure 3:**
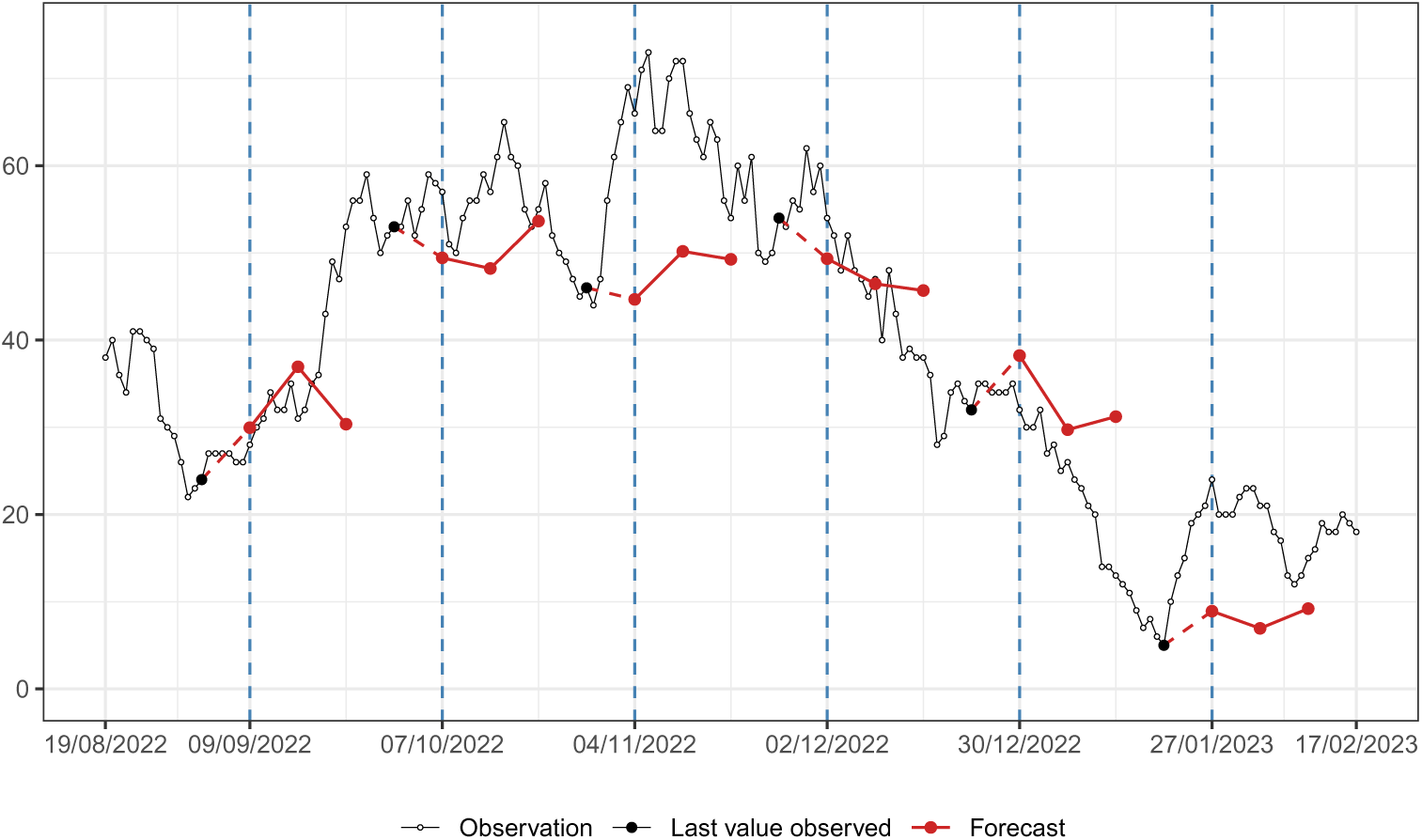
Examples of forecasts of weekly COVID-19-related hospital admissions to three weeks ahead during autumn 2022. Empty dots correspond to the number of COVID-19-related hospital admissions in the next seven days. For six dates set at regular intervals of four weeks, forecasts are generated using XGBoost for each of the following three weeks, based on COVID-19-related hospital admissions of the last 28 days.

Overall, all models except LR outperformed baseline when trained exclusively on counts of past COVID-19-related hospital admissions (feature set A), as assessed by both the summary score and the proportion of forecasts with lower RMSE than baseline (Table 2). The reductions in summary score were relatively modest (0.91 for RNN, 0.90 for LSTM and 0.88 for XGBoost), with XGBoost achieving the lower summary score and the most consistent performance across all combinations of lookback window, forecasting horizon, and train-test split (outperforming baseline in 76% of cases). With the exception of XGBoost, which maintained stable performance, all models performed worse on average on the partial study period compared to the full study period (Table 3). This decline in performance was even more pronounced for LR and LSTM model.

Including additional features beyond past COVID-19-related hospital admissions did not lead to any substantial improvement in the average summary score for any model on the full study period. For all feature sets except the number of COVID-19-related hospital admissions combined with the number of patients seeking care at the emergency ward (feature set C), XGBoost consistently outperformed both the baseline and all other models on the full study period (Table 2). RNN and LSTM performed similarly to baseline, only showing noticeable improvement when trained on the number of COVID-19-related hospital admissions alone (feature set A) or in combination with the number of patients seeking care at the emergency ward (feature set C). There was no feature set that enabled LR to produce more accurate forecasts than the baseline. The performance of LR was particularly poor when multiple features were added.

Adding measurements of SARS-CoV-2 viral load in wastewater to the feature set led to noticeable improvement of the performance of XGBoost on the partial study period (summary score 0.73 and improvement over baseline in 97% of the cases, Table 3). Using other feature sets, XGBoost led to slightly more precise forecasts on the partial study compared to the full study period. On the contrary, the other models generally performed worse on the partial study period than on the full study period. This drop in accuracy could be substantial, for instance for forecasts generated with LSTM using past COVID-19-related hospital admissions combined with counts of inpatients with high CRP value (feature set H, average summary score of 1.51 compared to 0.96).

Forecasting performance was highly dependent on forecasting horizon *k* and lookback window *p*, with the ranking of models and feature sets varying across the values chosen for *k* and *p*. At least one model outperformed the baseline for all combinations of *k* and *p*, in both the full and the partial study periods (Figure 4A). XGBoost was the best-performing model for 12 out of 25 combinations of *k* and *p* for the full study period (summary scores ranging from 0.67 to 0.84) and for 22 out of 25 combinations of *k* and *p* during the partial study period (summary scores ranging from 0.53 to 0.89). For smaller values of *k* and *p*, the RNN and LSTM models outperformed the other models, particularly for the full study period (summary scores ranging from 0.72 to 0.95). As the forecasting horizon and lookback window increased, the XGBoost model more frequently achieved the best performance. This pattern was more pronounced for the partial study period than for the full study period.

**Figure 4:**
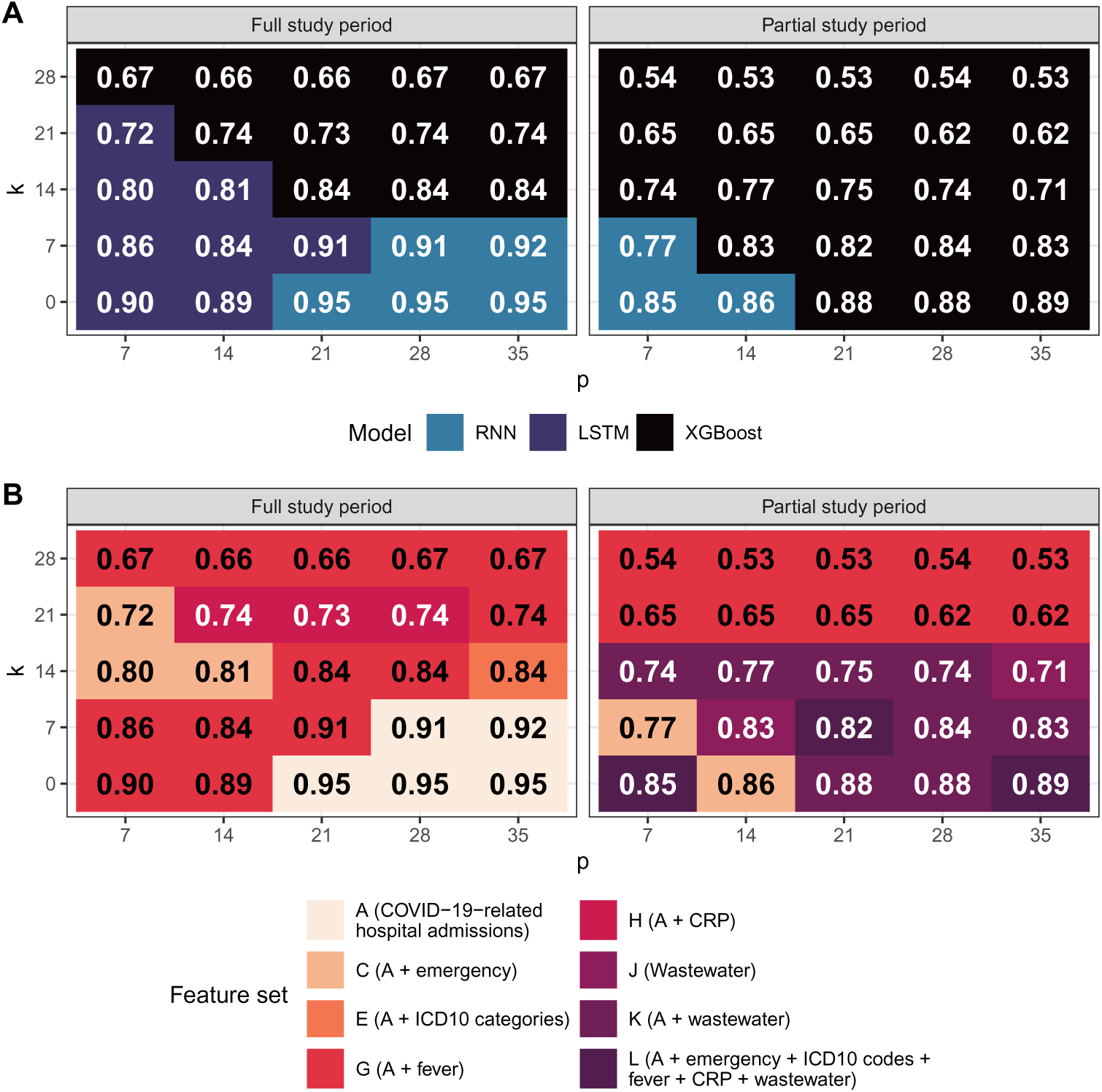
Best-performing model and feature set for each combination of forecasting horizon *k* and lookback window *p*. A: Model achieving the lowest root mean squared error (RMSE) during the full and the partial study period. B: Feature set achieving lowest RMSE during the full and the partial study period. Numbers indicate the average summary score, computed as the geometric mean of the ratios of RMSE over the baseline RMSE across all train-test splits.

The optimal feature set also varied according to experimental conditions. For 20 out of 25 combinations of *k* and *p* during the full study period and all 25 combinations of *k* and *p* during the partial study period, best performance was obtained using a feature set that included additional features besides COVID-19-related hospital admissions (Figure 4B). For the full study period, and for longer horizons for the partial study period, past COVID-19-related hospital admissions combined with counts of patients admitted with fever (feature set G) most frequently achieved the best summary score (summary score ranging from 0.53 to 0.91). Using counts of patients admitted with high CRP (feature set H) and counts of patients seeking care at the emergency ward (feature set C) were also sometimes selected as achieving best performance, especially for longer forecast horizons (three to five weeks ahead). The inclusion of viral load measurements in wastewater samples led to the best results at short forecast horizons (up to three weeks ahead) for the partial study period (summary score ranging from 0.71 to 0.88).

## 4. Discussion

In this study, we systematically evaluated and compared the ability of various machine learning algorithms to forecast the number of weekly COVID-19 hospital admissions up to five weeks ahead, using different combinations of variables extracted from EHR from six hospitals in the region of Bern, Switzerland, as well measurements of SARS-CoV-2 viral load in wastewater samples. Across all examined forecasting horizons, we were able to generate forecasts that consistently outperformed the baseline model of LOCF, with greater improvements observed for longer forecasting horizons. Overall, our findings confirm that EHR hold considerable potential for improving the forecasting of infectious disease dynamics.

We found that gradient boosting using the XGBoost algorithm outperformed other models on average across all combinations of forecasting horizon *k* and lookback window *p*. This is somewhat surprising as XGBoost was not inherently built for time series forecasting. Still, XGBoost performed better than linear regression and neural networks (RNN and LSTM), particularly for longer forecasting horizons. These findings may be explained by the discrete tree-based approach of XGBoost, leading to a good handling of non-linearities in addition to the reduced risk of overfitting (Park and Ho, 2021). Moreover, XGBoost may have an advantage because of the relative scarcity of data. As we work with daily time series, our models never get a training set containing more than about 1, 200 data points, which was further reduced when using longer lookback windows or focusing on the partial study period. In these situations, forecasts generated with linear regression and neural network were prone to perform considerably worse than LOCF, while XGBoost remained mostly adequate. This feature makes XGBoost particularly appealing in the early stages of epidemics and for emerging infectious diseases lacking historical data.

With regards to variables relevant for forecasting COVID-19-related hospital admissions, our findings indicate that relying solely on past admission counts is suboptimal. Complementing these data with additional variables available in EHR such as the number of patients admitted with fever, with elevated CRP or presenting to emergency care improved forecast performance, particularly at longer forecasting horizons (three to five weeks ahead). Besides EHR, our results confirm the transformative potential of incorporating viral load measurements in wastewater in infectious disease forecasting (Rankin et al.). Forecasts based on recent wastewater data demonstrated substantially improved performance for shorter horizons (up to three weeks ahead), while EHR-based variables such as fever-related admissions retained a performance advantage at longer horizons (four and five weeks ahead). This pattern likely reflects the temporal lag between infection incidence (captured by wastewater surveillance via fecal shedding) and subsequent hospital admissions, which has been estimated to range between 10 and 14 days (Hegazy et al., 2022).

Other studies have used similar approaches for hospital admission forecasting, and found that combining hospital admissions data on the level of a single hospital or aggregated on a regional level with additional health data (e.g., occupancy of emergency units or use of ambulance services) or external data (e.g., mobility or weather data) lead to more accurate forecasts (Ferté et al., 2022; Zhang et al., 2022; Klein et al., 2023). Our results are aligned with their findings, but a quantitative comparison of the precision of the obtained forecasts between studies is difficult due to different available data, study periods and evaluation metrics.

The main strength of our work lies in the breadth and thoroughness of the systematic comparison of different models and combinations of features, which leads to a proof-of-concept that routinely collected EHR can indeed provide a solid data basis for an infectious disease forecasting system. This represents a step forward in the development of infectious disease monitoring and forecasting systems relying on data that has not been collected specifically for research purposes. Given that the data can be accessed with little time delays, forecasting could be conducted continuously and provide reliable estimates of quantities of interest such as new COVID-19-related hospital admissions without depending on time-consuming and expensive data collection.

Our study comes with several limitations. First, the generalizability of our findings beyond the Insel Gruppe hospital network in the region of Bern remains uncertain. Differences in EHR structures, conventions and formats could make it difficult to replicate our study in other settings. We refrained from requesting access to EHR from other Swiss university hospitals. Second, we carried out a purely retrospective analysis, and did not implement our forecasting framework in a real-time operational context. Real-time deployment would require additional development of data pipelines and infrastructure. One key obstacle, which we could not influence, was the time lag between hospital admission and the encoding of diagnoses using ICD10 codes, which can occur several weeks after discharge. Reducing these delays is essential for enabling the practical application of forecasting approaches such as ours, although as we showed the best-performing features (fever, CRP and emergency ward) do not rely on ICD-10 encoding and are available immediately. Third, from a technical perspective, we did not use a distinct validation set to tune model hyperparameters, instead doing this directly on the testing set. This decision was made in light of limited data availability, as reserving additional data for validation would have reduced the training set. Similarly, we also did not estimate the uncertainty of the model forecasts. While techniques such as conformal prediction were considered, their application would have required additional data splitting, further reducing the training set. Finally, as with many forecasting approaches, we did not account for changes in transmission dynamics, for example due to shifts in population behavior, the emergence of new variants or increases in the immunity level due to vaccination. Future work is needed to develop forecasting methods that can incorporate a broader range of dynamic data sources.

The vast amount of routinely collected medical data remains underutilized for infectious disease forecasting. Our findings demonstrate that such data, when properly harnessed with modern machine learning approaches, can substantially enhance the accuracy of short-term hospital admission forecasts. Such forecasts are especially valuable for informing public health policy, enabling healthcare systems to anticipate surges in demand and allocate resources accordingly. As data infrastructures continue to expand, with more and more hospital data becoming available in standardized format and with decreasing delays, the integration of routine clinical and surveillance data into real-time forecasting systems will become more feasible. This paves the way for highly-efficient forecasting tools that can support timely and data-driven responses to emerging infectious disease threats, strengthening overall pandemic preparedness.

## Data Availability

Some data files are publicly available online on GitHub. Due to data protection regulations we can not make the full hospital dataset publicly available, but only in aggregated form.

https://github.com/mwohlfender/hospital_admission_forecasting

## Acknowledgments

This study is funded by the Multidisciplinary Center for Infectious Diseases, University of Bern, Bern, Switzerland. We gratefully acknowledge the Insel Data Science Center (IDSC) (www.idsc.io/en/) for facilitating the access to the electronic health records from the Insel Gruppe hospital network. Calculations were performed on UBELIX (www.id.unibe.ch/hpc), the HPC cluster at the University of Bern.

## Appendix A. Supplementary material

Detailed model description, table of hyperparameters, additional results and figures supporting the main text.

