## Appendix A. Supplementary material for "Machine learning-based short-term forecasting of COVID-19 hospital admissions using routine hospital patient data"

---

[*] Corresponding author

### 1. Model description

In this section we describe the structure of our forecasting models in detail using more formal notation than in the manuscript.

#### 1.1. Definitions and notation

We denoted the daily time series of the number of COVID-19 hospital admissions, i.e., the number of patients being assigned the ICD10 code U07.1 per day, by

$$X = \Big(x(t)\Big)_{t \geq 1}.$$

As day 1 we took 25 February 2020, the day the first COVID-19 case was confirmed in Switzerland (FOPH, Federal Office of Public Health, 2020). We had access to electronic health records until autumn 2023. Due to delays in recording the diagnoses of patients discharged from hospital in the form of ICD10 codes, we removed a few weeks of data at the end. The last day from which we included data into our models was therefore 30 June 2023. Consequently, our study period was the time between 25 February 2020 and 30 June 2023 and we could make the definition of $X$ more precise:

$$X = \Big(x(t)\Big)_{1 \leq t \leq T}, \quad \text{where} \quad T = 1222 \quad \text{corresponds to 30 June 2023.}$$

Since the wastewater data were only available starting from autumn 2021, we separately carried out our analysis on a second shortened study period, referred to as "partial study period", between 16 November 2021 and 30 June 2023. In order to distinguish the two study periods, we used the term "full study period" to refer to the original study period between 25 February 2020 and 30 June 2023.

#### 1.2. Features and target variable

We aimed to forecast the number of COVID-19 hospital admissions $k$ days ahead during a time interval of $N$ days. We defined

$$Y_{k,N} = \Big(y_{k,N}(t)\Big)_{1 \leq t \leq T-(k+N-1)}, \quad \text{where} \quad y_{k,N}(t) = \sum_{i=t+k}^{t+k+N-1} x(i).$$

Our focus lied on the cases

$$k = 0,\ 7,\ 14,\ 21 \text{ and } 28 \quad \text{and} \quad N = 7.$$

Some remarks:

(i) Setting $k = 0$ and $N = 7$ corresponds to $Y_{k,N}$ designating the number of COVID-19 hospital admissions during the upcoming week, i.e. during days $d$ to $d + 6$, where day $d$ corresponds to the current day.

(ii) Setting $k = 7$ and $N = 7$ corresponds to $Y_{k,N}$ designating the number of COVID-19 hospital admissions during a seven day time interval that starts seven days ahead, i.e. during days $d + 7$ to $d + 13$, where day $d$ corresponds to the current day.

As data basis for our models to forecast the time series $Y_{k,N}$, we used past values of daily time series, e.g., the number of patients seeking care at the emergency department of Bern University Hospital and the number of inpatients admitted to hospital with fever. We denoted these time series by

$$Z_1 = \Big(z_1(t)\Big)_{1\leq t\leq T}, \quad Z_2 = \Big(z_2(t)\Big)_{1\leq t\leq T}, \; \dots \;, \; Z_M = \Big(z_M(t)\Big)_{1\leq t\leq T}.$$

We used lagged values of $Z_1,\ Z_2,\ \dots,\ Z_M$ as features. More precisely, we aimed to estimate $y_{k,N}(t)$ based on

$$\begin{aligned}
&z_1(t-1),\ z_1(t-2),\ \dots,\ z_1(t-p),\\
&z_2(t-1),\ z_2(t-2),\ \dots,\ z_2(t-p),\\
&\dots\\
&z_M(t-1),\ z_M(t-2),\ \dots,\ z_M(t-p),
\end{aligned}$$

where $p$ denotes the lookback window, i.e., the number of past days of which data is used. In our analysis we focused on the cases

$$p = 7,\ 14,\ 21,\ 28 \text{ and } 35.$$

*1.3. Train-test splits*

We followed a standard machine learning workflow, fitting our models on a training dataset and evaluating our models on a separate testing dataset that does not intersect the training dataset. As all our features as well as our target variable were extracted from daily time series, we utilized this temporal structure when defining training and testing dataset. We used multiple splitting dates to divide the study periods into multiple subperiods for which forecasts shall be made (see Supplementary Figure S 1). We referred to these subperiods as "testing periods". More formally, each of these testing periods had the form

$$\left[\tau_{\text{min}}^{\text{test}},\ \tau_{\text{max}}^{\text{test}}\right],$$

where $\tau_{\text{min}}^{\text{test}}$ is one of the splitting dates and $\tau_{\text{max}}^{\text{test}}$ is one day before the subsequent splitting date. We used all instances of the features that were available on the day $\tau_{\text{min}}^{\text{test}}$ for training the models, i.e., all values of the features observed during the study period no later than the day before $\tau_{\text{min}}^{\text{test}}$.

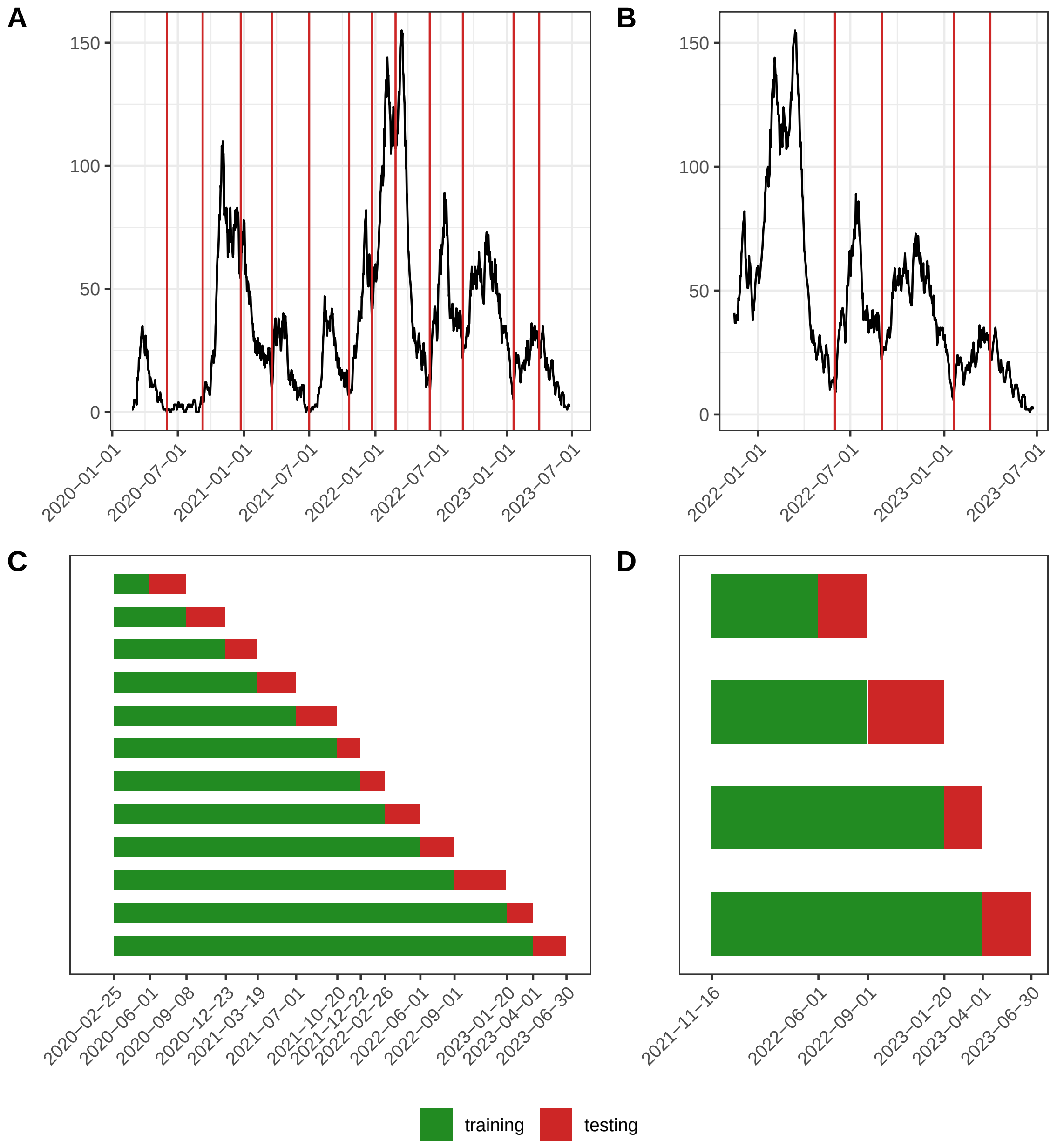

Supplementary Figure S 1: **COVID-19 hospital admissions and train-test splits.**
A and B: Number of COVID-19 hospital admissions in next seven days (days $t$ to $t + 6$) and splitting dates to define testing periods for full and partial study period.
C and D: Periods during which the data used for training was collected and periods for which forecasts of COVID-19 hospital admissions shall be made.

*1.4. Feature matrix and target variable vector*

We put the feature matrix and target variable vector of the testing dataset into chronological form. Each row corresponds to one day of the testing period. A row of the feature matrix contains values of the time series $Z_1$, $Z_2$, ..., $Z_M$ collected 1 to $p$ days before the corresponding day of the testing period. Accordingly, an entry of the target variable vector contains the number of COVID-19-related hospital admissions during a time interval of $N$ days starting $k$ days ahead of the corresponding day of the testing period.

Table 1: Feature matrix and target variable vector of testing dataset.

| Day | Features: Past values of $Z_1$, ... , $Z_M$ | | | Target variable |
|---|---|---|---|---|
| $t$ | $z_1(t-1)$ | ... | $z_M(t-p)$ | $y_{k,N}(t)$ |
| $\tau_{\min}^{\text{test}}$ | $z_1(\tau_{\min}^{\text{test}}-1)$ | ... | $z_M(\tau_{\min}^{\text{test}}-p)$ | $y_{k,N}(\tau_{\min}^{\text{test}})$ |
| $\tau_{\min}^{\text{test}}+1$ | $z_1(\tau_{\min}^{\text{test}})$ | ... | $z_M(\tau_{\min}^{\text{test}}+1-p)$ | $y_{k,N}(\tau_{\min}^{\text{test}}+1)$ |
| $\tau_{\min}^{\text{test}}+2$ | $z_1(\tau_{\min}^{\text{test}}+1)$ | ... | $z_M(\tau_{\min}^{\text{test}}+2-p)$ | $y_{k,N}(\tau_{\min}^{\text{test}}+2)$ |
| ... | ... | ... | ... | ... |
| $\tau_{\max}^{\text{test}}$ | $z_1(\tau_{\max}^{\text{test}}-1)$ | ... | $z_M(\tau_{\max}^{\text{test}}-p)$ | $y_{k,N}(\tau_{\max}^{\text{test}})$ |

We defined the training dataset in dependence on the testing period. All data included into the feature matrix or the target variable vector of the training dataset was collected before $\tau_{\min}^{\text{test}}$. The structure of feature matrix and target variable vector of the training dataset is the same as for the testing dataset. We used the notation

$$\left[\tau_{\min}^{\text{train}},\ \tau_{\max}^{\text{train}}\right]$$

to denote the days corresponding to the rows of the feature matrix and target variable vector of the training dataset and referred to it as "training period".

Table 2: Feature matrix and target variable vector of training dataset.

| Day | Features: Past values of $Z_1$, ... , $Z_M$ | | | Target variable |
|---|---|---|---|---|
| $t$ | $z_1(t-1)$ | ... | $z_M(t-p)$ | $y_{k,N}(t)$ |
| $\tau_{\min}^{\text{train}}$ | $z_1(\tau_{\min}^{\text{train}}-1)$ | ... | $z_M(\tau_{\min}^{\text{train}}-p)$ | $y_{k,N}(\tau_{\min}^{\text{train}})$ |
| $\tau_{\min}^{\text{train}}+1$ | $z_1(\tau_{\min}^{\text{train}})$ | ... | $z_M(\tau_{\min}^{\text{train}}+1-p)$ | $y_{k,N}(\tau_{\min}^{\text{train}}+1)$ |
| $\tau_{\min}^{\text{train}}+2$ | $z_1(\tau_{\min}^{\text{train}}+1)$ | ... | $z_M(\tau_{\min}^{\text{train}}+2-p)$ | $y_{k,N}(\tau_{\min}^{\text{train}}+2)$ |
| ... | ... | ... | ... | ... |
| $\tau_{\max}^{\text{train}}$ | $z_1(\tau_{\max}^{\text{test}}-1)$ | ... | $z_M(\tau_{\max}^{\text{test}}-p)$ | $y_{k,N}(\tau_{\max}^{\text{train}})$ |

Some properties of $\tau_{\min}^{\text{train}}$ and $\tau_{\max}^{\text{train}}$:

(i) The lowest possible value for $\tau_{\min}^{\text{train}}$ is

$$\tau_{\min}^{\text{train}} = p + 1$$

because each row of the feature matrix of the training dataset contains lagged values of $Z_1$, ... , $Z_M$ of the previous $p$ days.

(ii) The most reasonable choice for $\tau_{\max}^{\text{train}}$ is

$$\tau_{\max}^{\text{train}} = \tau_{\min}^{\text{test}} - (k + N)$$

because

$$y_{k,N}\big(\tau^{\text{test}}_{\min} - (k+N)\big) = x(\tau^{\text{test}}_{\min} - N) + x(\tau^{\text{test}}_{\min} - N + 1) + ... + x(\tau^{\text{test}}_{\min} - 1)$$

contains data of day $\tau^{\text{test}}_{\min} - 1$, the last day of which data is available on day $\tau^{\text{test}}_{\min}$. Therefore, no additional complete row of more recent data can be added to the feature matrix and target variable vector of the training dataset.

(iii) Forecasting $Y_{k,N}$ in a forecasting setup defined by the parameters $k$, $N$ and $p$, training period

$$\big[\tau^{\text{train}}_{\min},\ \tau^{\text{test}}_{\min} - (k+N)\big]$$

and testing period

$$\big[\tau^{\text{test}}_{\min},\ \tau^{\text{test}}_{\max}\big]$$

requires data of the period $\big[\tau^{\text{train}}_{\min} - p,\ \tau^{\text{test}}_{\max} + k + N - 1\big]$.

*1.5. Definition of summary score*

We developed a summary metric to assess the impact of model, feature set and parameters $k$ and $p$ on the average precision of the forecasts across train-test splits of the study period. In this paragraph we present the definition of this summary score using the formal language introduced in the previous paragraphs.

Let $\hat{y}_1, \hat{y}_2, \dots, \hat{y}_n$ be a collection of forecasts. Each $\hat{y}_i$ is the forecast for the testing period $\big[\tau^{\text{test}}_{\text{min, i}},\ \tau^{\text{test}}_{\text{max, i}}\big]$ obtained by the model $M_i$, based on the feature set $F_i$ and depending on the parameters $k_i$, $N_i$ and $p_i$. It has the following form:

$$\hat{y}_i = \left(\hat{y}_i\left(\tau^{\text{test}}_{\text{min, i}}\right),\ \hat{y}_i\left(\tau^{\text{test}}_{\text{min, i}} + 1\right), \dots,\ \hat{y}_i\left(\tau^{\text{test}}_{\text{max, i}}\right)\right).$$

The summary score is computed as follows:

$$\Big(\prod_{i=1}^{n} \frac{\text{RMSE}\big(\hat{y}_i, y_{k_i,N_i}\big)}{\text{RMSE}\big(\tilde{y}_i, y_{k_i,N_i}\big)}\Big)^{\frac{1}{n}},$$

where

$$\tilde{y}_i = \left(y_{0,N_i}\left(\tau^{\text{test}}_{\text{min, i}} - N_i\right),\ y_{0,N_i}\left(\tau^{\text{test}}_{\text{min, i}} - N_i + 1\right), \dots,\ y_{0,N_i}\left(\tau^{\text{test}}_{\text{max, i}} - N_i\right)\right)$$

is the forecast of the baseline model last observation carried forward (LOCF) for the testing period $\big[\tau^{\text{test}}_{\text{min, i}},\ \tau^{\text{test}}_{\text{max, i}}\big]$,

$$\text{RMSE}\big(\hat{y}_i, y_{k_i,N_i}\big) = \sqrt{\frac{1}{D_i}\sum_{j=0}^{D_i-1}\Big(\hat{y}_i\left(\tau^{\text{test}}_{\text{min, i}} + j\right) - y_{k_i,N_i}\left(\tau^{\text{test}}_{\text{min, i}} + j\right)\Big)^2}$$

and

$$\begin{aligned}\mathrm{RMSE}\big(\tilde{y}_i, y_{k_i,N_i}\big) &= \sqrt{\frac{1}{D_i}\sum_{j=0}^{D_i-1}\Big(\tilde{y}_i\left(\tau^{\text{test}}_{\text{min, i}}+j\right) - y_{k_i,N_i}\left(\tau^{\text{test}}_{\text{min, i}}+j\right)\Big)^2} \\ &= \sqrt{\frac{1}{D_i}\sum_{j=0}^{D_i-1}\Big(y_{0,N_i}\left(\tau^{\text{test}}_{\text{min, i}}-N_i+j\right) - y_{k_i,N_i}\left(\tau^{\text{test}}_{\text{min, i}}+j\right)\Big)^2}\,,\end{aligned}$$

where $D_i$ is the length of the testing period $\left[\tau^{\text{test}}_{\text{min, i}},\ \tau^{\text{test}}_{\text{max, i}}\right]$.

$\mathrm{RMSE}\big(\hat{y}_i, y_{k_i,N_i}\big)$ is the root mean square error between the forecast of the model $M_i$ and the observations and $\mathrm{RMSE}\big(\tilde{y}_i, y_{k_i,N_i}\big)$ is the root mean square error between the forecast of the baseline model LOCF and the observations.

### 2. Hyperparameters

To find the optimal configuration of our machine learning models, we ran the recurrent neural network (RNN) model, the long short-term memory (LSTM) model and the XGBoost model with different values of their respective hyperparameters.

Supplementary Table S 1: **Range of values of hyperparameters.** The models were run with all possible combinations of the values of the hyperparameters listed in this table.

| Model | Combinations | Parameter | Values |
|---|---|---|---|
| RNN | 144 | number of hidden layers | 1, 2 |
| | | neurons in first hidden layer | 16, 32, 64 |
| | | neurons in second hidden layer | 8, 16, 32 |
| | | activation function | relu |
| | | optimizer | adam |
| | | learning rate | 0.0005, 0.001, 0.002, 0.005 |
| | | number of epochs | 10, 25, 50 |
| LSTM | 144 | number of hidden layers | 1, 2 |
| | | neurons in first hidden layer | 16, 32, 64 |
| | | neurons in second hidden layer | 8, 16, 32 |
| | | activation function | relu |
| | | optimizer | adam |
| | | learning rate | 0.0005, 0.001, 0.002, 0.005 |
| | | number of epochs | 10, 50, 100 |
| XGBoost | 864 | maximum number of boosting iterations | 10, 25, 50, 100, 150, 200 |
| | | learning rate (eta) | 0.1, 0.3, 0.5, 0.7 |
| | | maximum depth of a tree | 3, 4, 5, 6, 7, 8 |
| | | maximum change allowed between iterations (max delta step) | 0, 5, 10, 15, 20, 25 |

### 3. Additional results

In this section we present additional figures showing how well the different machine learning models did in forecasting COVID-19 hospital admissions, both during the full and during the partial study period. First, we present the results displayed in Tables 2 and 3 in the manuscript in more detail. More precisely, for each model, each feature set and each combination of forecasting horizon $k$ and lookback window $p$ we display the summary score computed as geometric mean of ratios of root mean square error (RMSE) of the model by RMSE of the baseline model last observation carried forward (LOCF) between forecast and true observations across all train-test splits. Second, for each model and each feature set we compare the average accuracy of the obtained forecasts. We compute the summary score for each model and each combination of forecasting horizon $k$ and lookback window $p$ across all feature sets and all train-test splits and the summary score for each feature set and each combination of forecasting horizon $k$ and lookback window $p$ across all models and all train-test splits. Finally, we determine for each combination of forecasting horizon $k$ and lookback window $p$ which model achieved the lowest summary score across all feature sets and all train-test splits and which feature set lead to the lowest summary score across all models and all train-test splits.

#### *3.1. Full study period*

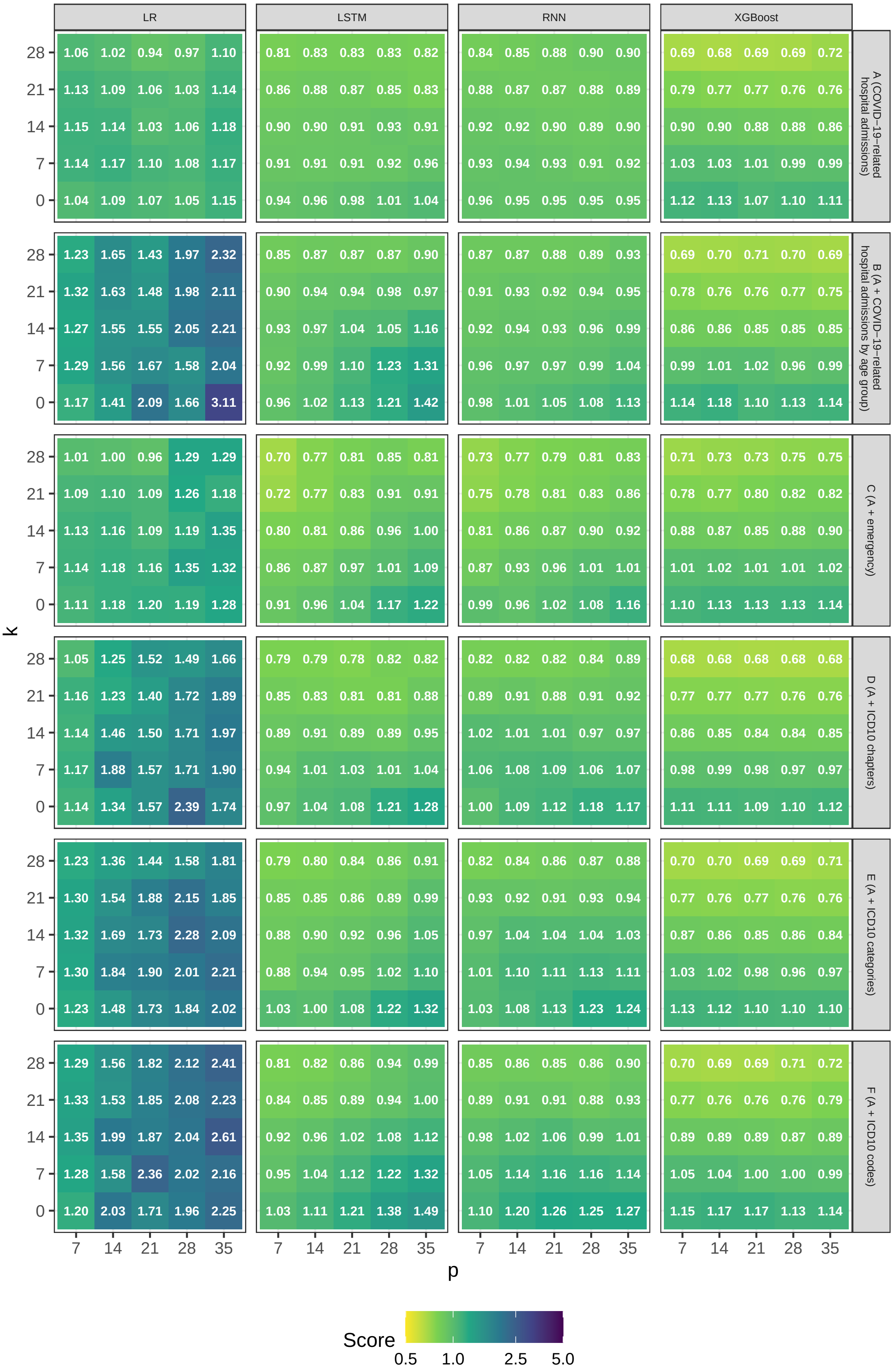

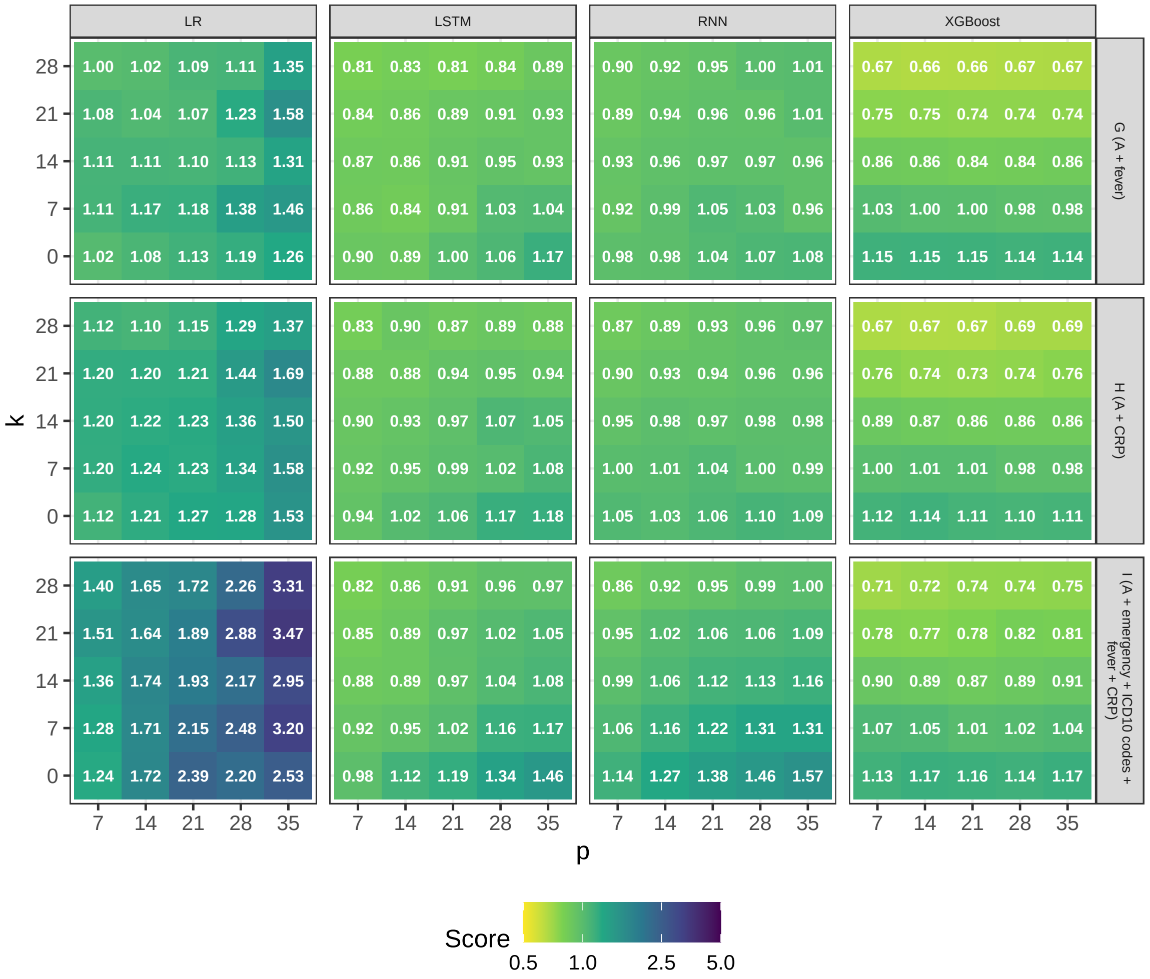

Supplementary Figure S 2: **Overview of accuracy of forecasts during full study period.** summary score computed as geometric mean of ratios of RMSE of model by RMSE of baseline model LOCF between forecast and true observations across all train-test splits of full study period.

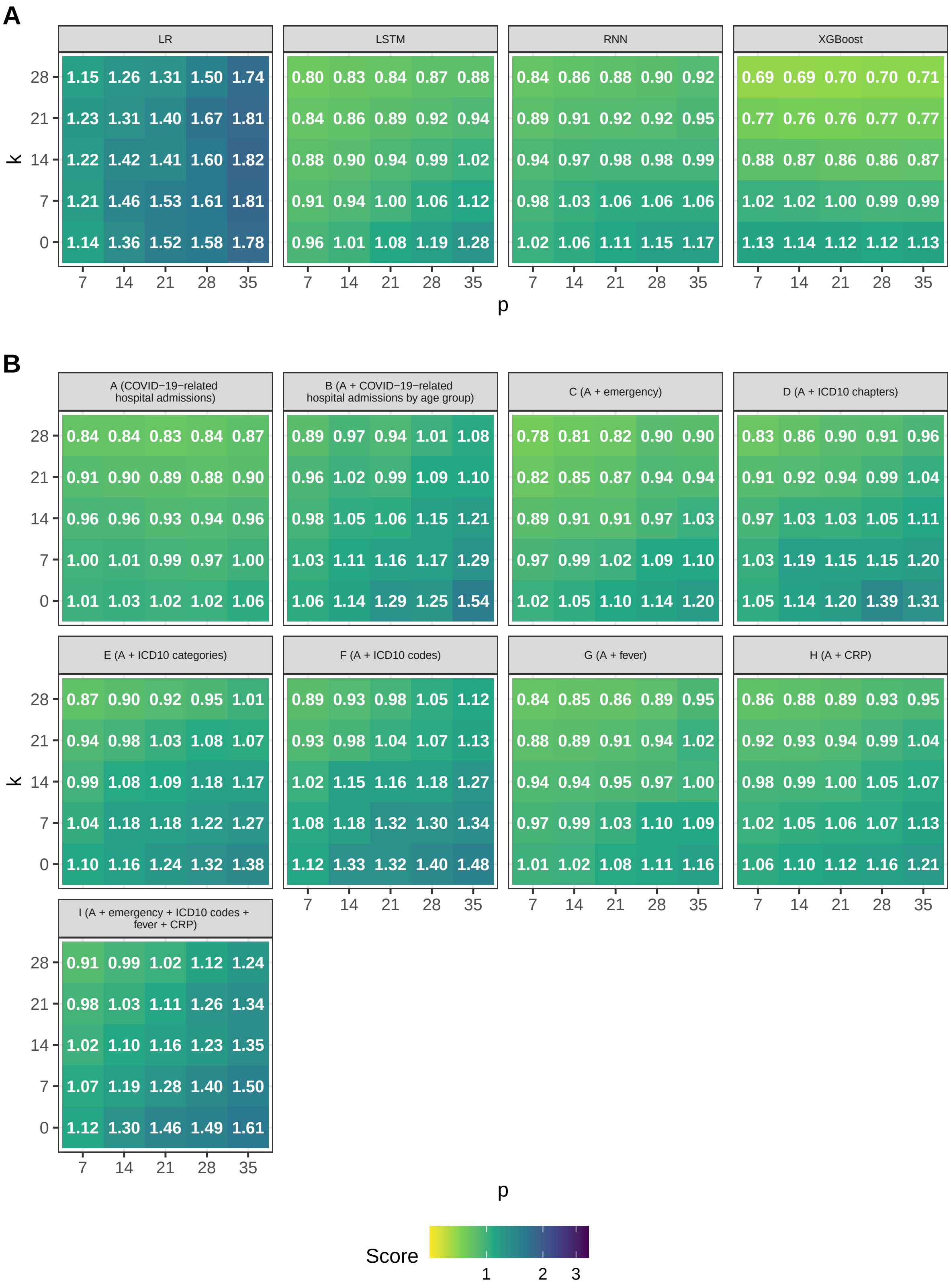

Supplementary Figure S 3: **Average accuracy of forecasts during full study period by model and by feature set.** A: summary score computed as geometric mean of ratios of RMSE of model by RMSE of baseline model LOCF between forecast and true observations across all feature sets and all train-test splits of full study period. B: summary score computed across all models and all train-test splits of full study period.

### *3.2. Partial study period*

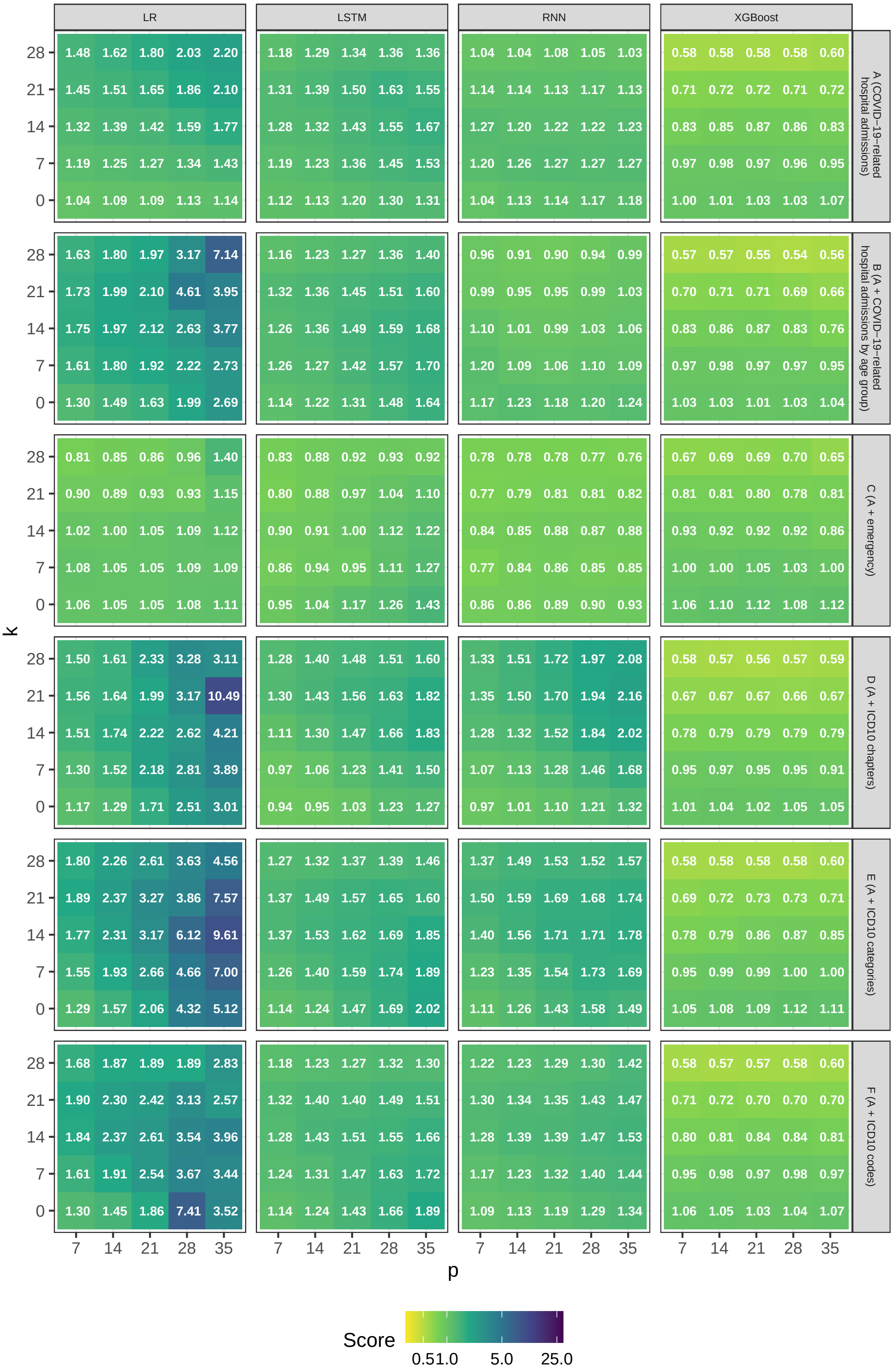

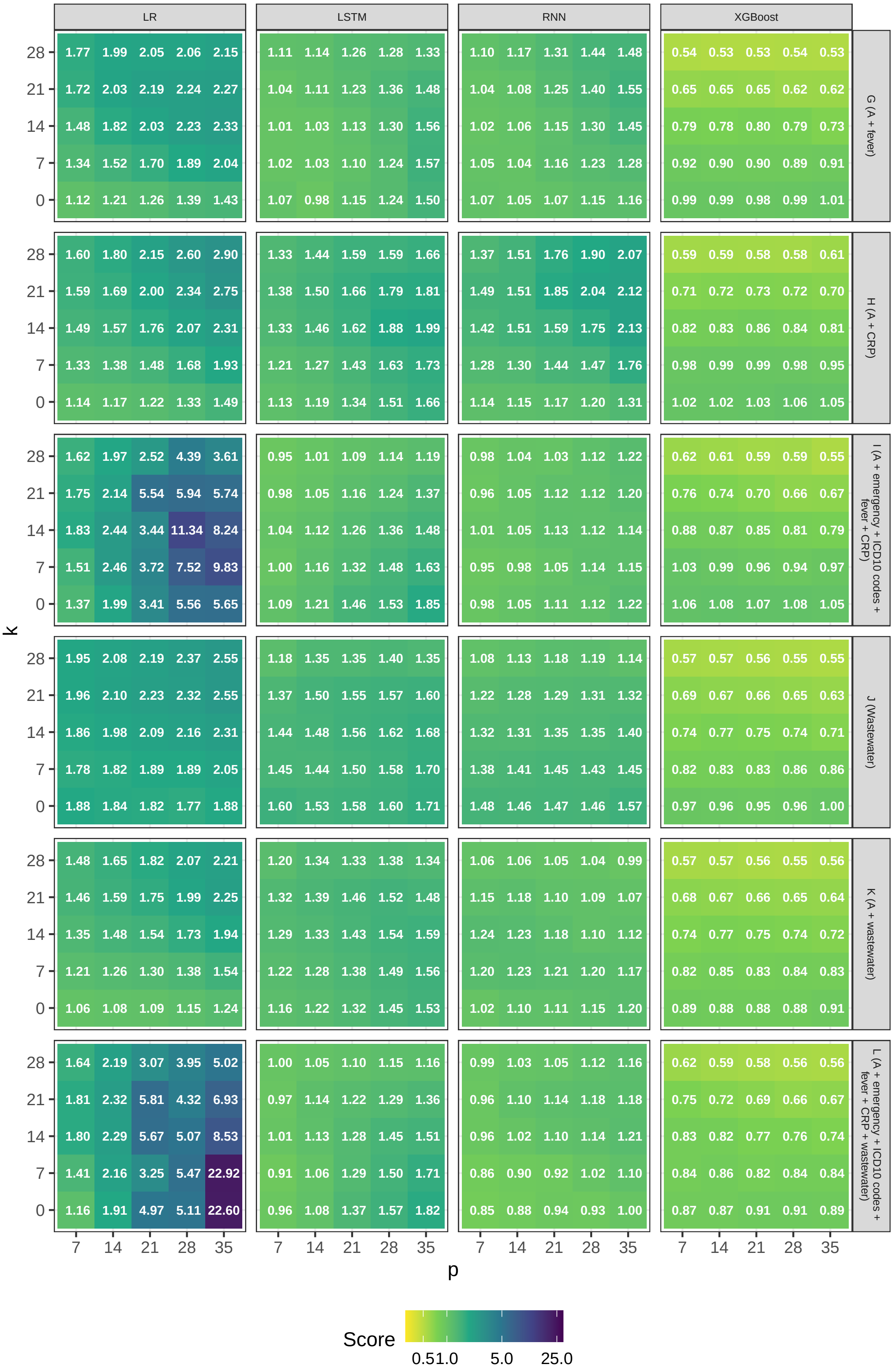

Supplementary Figure S 4: **Overview of accuracy of forecasts during partial study period.** summary score computed as geometric mean of ratios of RMSE of model by RMSE of baseline model LOCF between forecast and true observations across all train-test splits of partial study period.

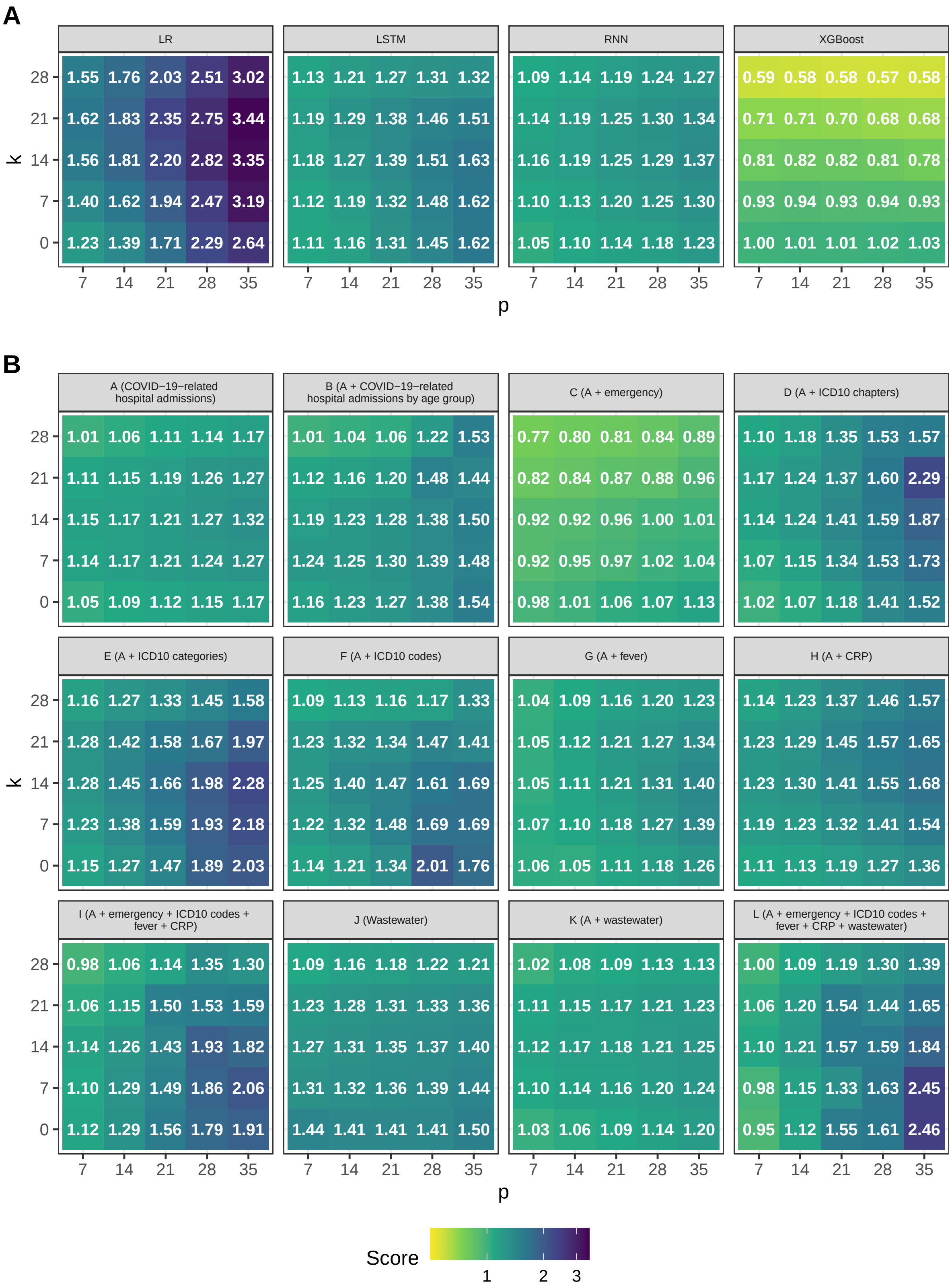

Supplementary Figure S 5: **Average accuracy of forecasts during partial study period by model and by feature set.** A: summary score computed as geometric mean of ratios of RMSE of model by RMSE of baseline model LOCF between forecast and true observations across all feature sets and all train-test splits of partial study period. B: summary score computed across all models and all train-test splits of partial study period.

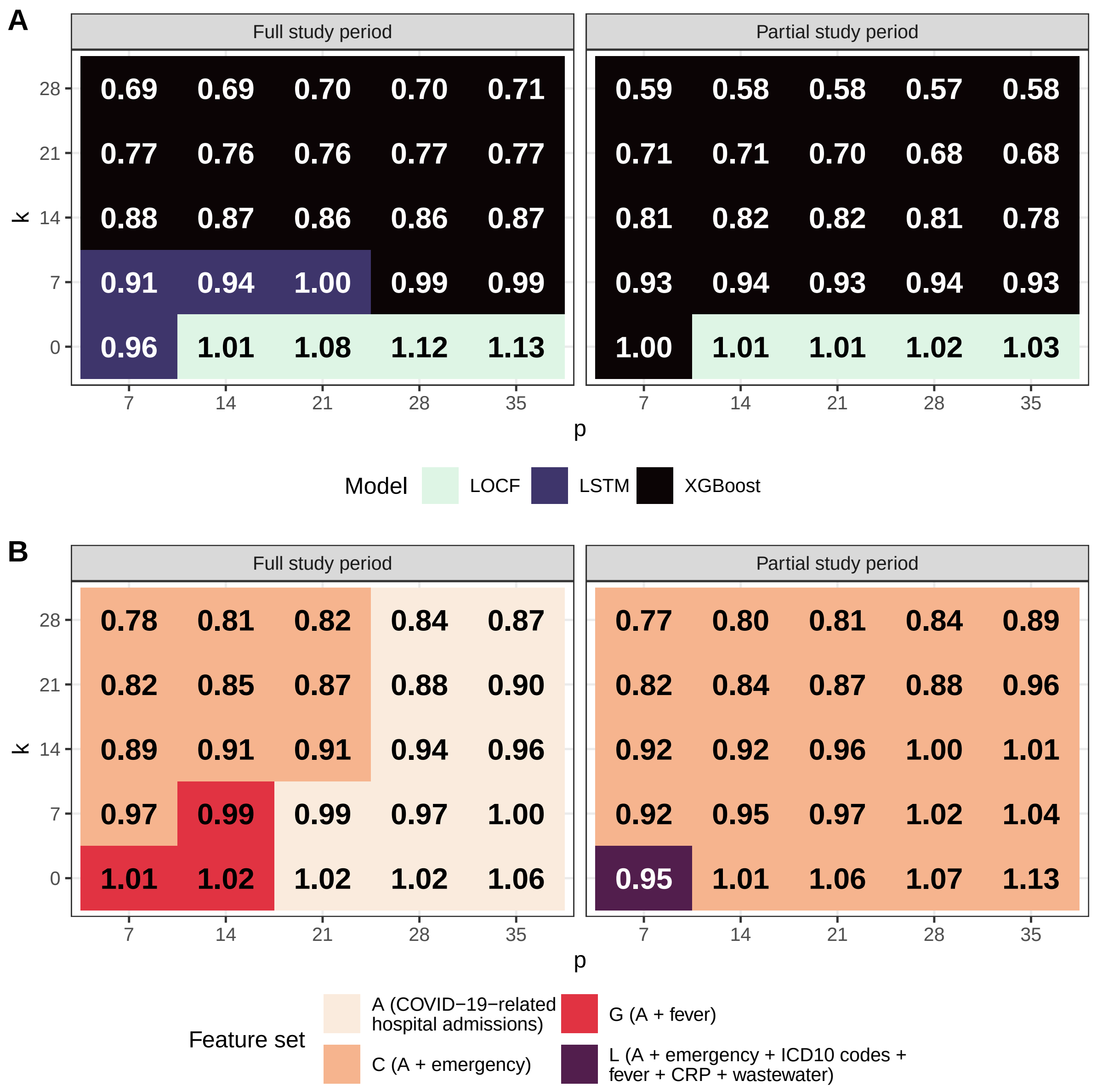

Supplementary Figure S 6: **Model and feature set producing forecasts with highest average accuracy.** A: Model achieving lowest summary score computed as geometric mean of ratios of RMSE of model by RMSE of baseline model LOCF between forecast and true observations across all feature sets and all train-test splits of full and partial study period for all combinations of forecasting horizon $k$ and lookback window $p$. B: Feature set leading to lowest summary score across all models and all train-test splits of full and partial study period for all combinations of forecasting horizon $k$ and lookback window $p$.
